# Vascular Thrombosis in COVID-19: A Potential Association with Antiphospholipid Antibodies

**DOI:** 10.1101/2020.11.02.20224642

**Authors:** Aneesh S Kallapur, Eric Y Yen, Ram Raj Singh

**Author notes:** **Corresponding author:** Ram Raj Singh, MD, Autoimmunity and Tolerance Laboratory, Division of Rheumatology, UCLA, 1000 Veteran Avenue, Room 32-59, Los Angeles, CA 90095-1670.

## Abstract

**Background:** Vascular thrombosis is common in patients with coronavirus disease 2019 (COVID-19). Etiologies underlying this complication are unclear.

**Purpose:** To determine the prevalence of antiphospholipid (aPL), including lupus anticoagulant, anti-cardiolipin and anti-β2-glycoprotein-1 antibodies, and its possible association with thrombotic manifestations of COVID-19.

**Data Sources:** We searched MEDLINE indexed journals on September 24, 2020 using the tool LitCovid and the pre-print server medRxIV.

**Study Selection:** Original investigations (cross-sectional studies, cohort studies, case series, and research letters) on COVID-19 and thrombosis were included.

**Data Extraction:** Data were independently extracted, and compiled into spreadsheets based on the PRISMA principles.

**Data Synthesis:** Hospitalized patients with COVID-19 showed a higher prevalence of lupus anticoagulant compared to non-COVID-19 patients. Temporally, lupus anticoagulant was generally positive early in the course of illness, whereas anti-cardiolipin and anti-β2-glycoprotein-1 antibodies appeared to emerge later in the disease. Some patients who were aPL-negative at an early time-point after disease onset became aPL-positive at a later time-point. Lupus anticoagulant was independently associated with thrombosis in 60 COVID-19 patients in New York had who had 32 thrombotic events (8 arterial and 24 venous). In 88 patients in Wuhan, who had more than 20 each of arterial and venous thrombotic events, medium/high positivity for multiple aPL was significantly associated with arterial thrombosis. However, the association of aPL with thrombosis was not evident in reports that had an overall lower number of or predominantly venous thrombotic events. Analysis of pooled patients revealed that aPL were significantly more frequent in COVID-19 patients with stroke than stroke patients in the general population. Furthermore, injection of IgG aPL fractions from COVID-19 patients into mice accelerated venous thrombosis.

**Limitation:** Limited data and paucity of prospective studies.

**Conclusion:** The aPL are prevalent in patients with COVID-19 and their presence is associated with thrombosis. Importantly, these antibodies may be a key mechanism of thrombosis in COVID-19. Follow-up studies are required to understand the relationship between aPL and the spectrum of vascular thrombosis during and after infection with SARS-CoV-2.

**Primary Funding Source:** None.

## INTRODUCTION

The coronavirus disease 2019 (COVID-19) pandemic, caused by the severe acute respiratory syndrome coronavirus-2 (SARS-CoV-2) has resulted in tremendous morbidity and mortality. Many patients with COVID-19 exhibit hypercoagulable states, with manifestations ranging from disseminated intravascular coagulation to venous thromboembolism and cerebrovascular disease (1). Among patients with acute respiratory distress syndrome, more thrombotic complications were diagnosed in COVID-19 patients than in matched patients without COVID-19 (odds ratio 2.6 [1.1–6.1]) (2). An autopsy study detected venous thromboembolism in more than half of deceased COVID-19 patients in whom it was not suspected earlier (3).

As these thrombotic manifestations began being reported at high rates, attempts are being made to uncover the underlying etiologies. A report from Wuhan, China, in April 2020 described three COVID-19 patients who developed thrombosis including multiple-vessel brain infarction and acro-ischemia (4). All three patients had anti-cardiolipin (aCL) and anti-β2 glycoprotein-1 (aβ2GP1) antibodies, collectively called antiphospholipid (aPL) antibodies.

The presence of aPL antibodies and a functional assay called lupus anticoagulant (LA) are the hallmarks of antiphospholipid syndrome (APS) that manifests with venous and arterial thrombosis (5). Some patients with APS have no other associated condition (primary APS), while others can be associated with infections and autoimmune disorders. Various infections, notably Hepatitis C virus (HCV) and HIV, have been shown to induce aPL antibodies, however, the significant thrombotic risk with aPL was seen only in HCV infection (6, 7). It is unclear whether SARS-CoV-2 elicits the production of aPL that associate with increased thrombotic risk.

In this systematic review, we identify patients with COVID-19 and thrombosis who were tested for aPL, and determine the prevalence of aPL in this population compared to the general population with reference to specific thrombotic manifestations. We further seek if any causal relationship exists between the presence of aPL in these patients and thrombosis.

## Methods

### Data Sources and Searches

We searched MEDLINE indexed journals using the tool LitCovid (8) on September 24, 2020. Due to the rapidly changing landscape of research in COVID-19, we also searched the pre-print server medRxIV, with the acknowledgment that those articles are not peer-reviewed.

### Study Selection

We included all original investigations (cross-sectional studies, cohort studies, case series, case reports and research letters) relevant to adult patients in the English language. We used the following search terms: “COVID-19”, “Coronavirus”, “ncov”, “2019-ncov” and “Antiphospholipid antibodies”, “Antiphospholipid antibody syndrome”, “Lupus anticoagulant” and “Stroke”, “Venous thromboembolism”, “Deep vein thrombosis” (DVT), “Myocardial infarction” and “Acute limb ischemia”. Articles that did not contain any laboratory data and those which did not carry out testing for aPL were excluded.

### Data Extraction and Quality Assessment

Data were extracted by one reviewer (A.K.), and independently reviewed by other two authors. We assessed article for the quality of evidence (**Appendix Table 1**).

**Table 1.** Prevalence of positive antiphospholipid assays in patients with COVID-19

| Location [Ref.] | Patient cohort | LA | aCL | aβ2GP1 | Comment |
| --- | --- | --- | --- | --- | --- |
| UK (9) | 44 (20%) of 216 patients with SARS-CoV-2 PCR+ vs. 43 (8%) of 540 control specimens sent for LA testing had a prolonged aPTT | Among those with a prolonged aPTT, LA+ in 31/34 (91%) patients with SARS-CoV-2 vs. 11/43 (26%) controls (p<0.001) | N/A | N/A | Heparin detected in most specimens, but the dRVVT assay contains heparinase, which neutralizes any heparin effect that might lead to false positive LA. |
| France (18) | 25 critically ill COVID-19 patients | 92% (dRVVT) | 52% | 12% | Single aPL 32%, double 52%, triple 12% |
| New York, USA (10) | All LA test orders at a tertiary hospital: 68 SARS-CoV-2+ and 119 negative | SARS-CoV-2+ patients: 44%, SARS-CoV-2-: 22% (p 0.002) | 1 IgM aCL+ among LA+ patients | 1 IgM aβ2GP1+ among LA+ patients | IgA antibodies not tested. Disease duration not known. |
| Italy (11) | 122 patients with COVID-19, 157 primary APS, and 91 oARD | COVID-19: 22%*<br>Primary APS: 54%<br>oARD: 15% | **IgG 13%**, IgM 3%, IgA 2%.<br>IgG 68%, IgM 38%.<br>IgG 13%, IgM 14%. | IgG 6%, IgM 7%, IgA 3%.<br>IgG 71%, IgM 43%.<br>IgG 21%, IgM 11%. | Frequency of aPL in COVID-19 lower vs. primary APS, but LA, IgG aCL, and IgM aβ2GP1 frequencies comparable to oARD. |
| France (41) | 56 patients with COVID-19 | 56% (dRVVT, LA-sensitive aPTT) | 10% (IgM/IgG aCL or | aβ2GP1) | IgA antibodies not tested. |
| Spain (19) | 24 patients with VTE admitted to ICU | NA | 8.3% with weakly + IgM | 8.3% with weakly + IgM | Tested within 14 days of admission |
| Italy (15) | 122 severe or critical patients | NA (Prolonged aPTT in 58%) | IgG 6%, IgM 7% | **IgG 16%**, IgM 9%, IgA 7% | aPS/PT IgG 3%, IgM 10% |
| Belgium (20) | 31 consecutive patients admitted to ICU | 67% | **IgG 19%**, IgM 3%, IgA 10% | **IgG 10%**, IgM 3%, IgA 10%. | 74% patients had at least 1 aPL.† aPS/PT: 3 IgG, 4 IgM. (All 7 LA+) |
| China (12) | Wuhan cohort: 66 critically ill, older (mean age ≥65 years) patients, Beijing cohort: 13 non-critically ill with a mean age of 35 years | Wuhan: 3% (dRVVT). 0/13 of Beijing cohort vs. 47% (31/66) Wuhan patients had an aPL. | IgG 6%, IgM 3%, **IgA 26%** (Wuhan cohort, n = 66). | IgG 18%, IgM 2%, **IgA 29%** (Wuhan cohort). | aPL emerged ~35-39 days post-disease onset. 10 aPL- patients at early time point became aPL+ at a later time point. Any aPL 47%, double+ 44%, triple+ 3% |
| China (13) | 19 critically ill patients admitted to ICU in Wuhan | 5% (dRVVT). 53% patients had at least 1 aPL. | IgG 11%, IgM 5%, IgA 32% | IgG 32%, IgM 0, IgA 37%. | Specimens obtained 29-32 days after symptom onset. ‡ |
| France (14) | 74 consecutive mechanically ventilated patients on anticoagulation | 85% (dRVVT). 88% patients had at least 1 aPL. | 12% (IgG/IgM aCL and/or IgG aβ2GP1). | 1 triple+, 2 LA- aCL/aβ2GP1+ | IgA antibodies and IgM aβ2GP1 not tested. # |
| Mexico (17) | 21 patients with severe or critical COVID-19 (18 anticoagulated) | NA | IgG 10%, IgM 14%. | IgG 5%, IgM 0. | Anti-annexin V: IgG 5%, IgM 19%<br>aPS: IgG 10%, IgM 14% |
| Michigan, USA (16) | 172 patients hospitalized with COVID-19. | NA. Any aPL + in 52%. | IgG 5%, IgM 23%, IgA 4% | IgG 3%, IgM 5%, IgA 4% | aPS/PT: IgG in 24%, IgM 18%.<br>>1 type of aPL in 24%, and >2 in 8%. ⊕ |

\* LA tested by dRVVT and silica clotting time. †2 patients triple+, 3 LA+IgG aCL+, 2 LA-IgG aCL+ and/or IgG aβ2GP1+. 9/10 LA+ patients tested negative on a 2<sup>nd</sup> occasion. ‡ Seven of 10 patients had multiple isotypes. 1 triple +. Some patients had very high levels of IgA antibodies. # aCL/aβ2GP1+ patients tended to have been
tested longer after COVID-19 symptoms started than the negative patients ( $p = .06$ ). ⊕ Moderate-to-high level aPL antibodies present in 30% patients. Most specimens collected at 0-15 hospital day.
+, positive; –, negative; aβ2GP1, anti-β2 glycoprotein 1 antibody; aCL, anti-cardiolipin antibody; aPL, antiphospholipid antibody; APS, antiphospholipid syndrome; aPS, anti-phosphatidylserine; aPT, anti-prothrombin; aPTT, activated partial-thromboplastin time; dRVVT, diluted Russell viper venom test; ICU, intensive care unit; LA, lupus anticoagulant; NA, not assessed / not available; oARD, other autoimmune rheumatic diseases.

### Data Synthesis and Analysis

First, to assess the prevalence of aPL in COVID-19, we compiled studies that reported aPL assays in patients with COVID-19. Second, we compiled studies that reported aPL assays in COVID-19 patients in relation to thrombosis. Third, we compared the frequency of aPL in patients with stroke in the general population versus COVID-19 using Fisher’s exact test.

## Results

We found 4,354 search results for these queries, of which 221 results were relevant (**Appendix Figure 1**). After excluding 59 articles that did not report any laboratory parameters and 104 articles that did not test for aPL antibodies, we were left with 58 articles reporting thrombotic events in COVID-19 patients who were tested for aPL antibodies, which we included in our systematic review.

**Figure 1:**
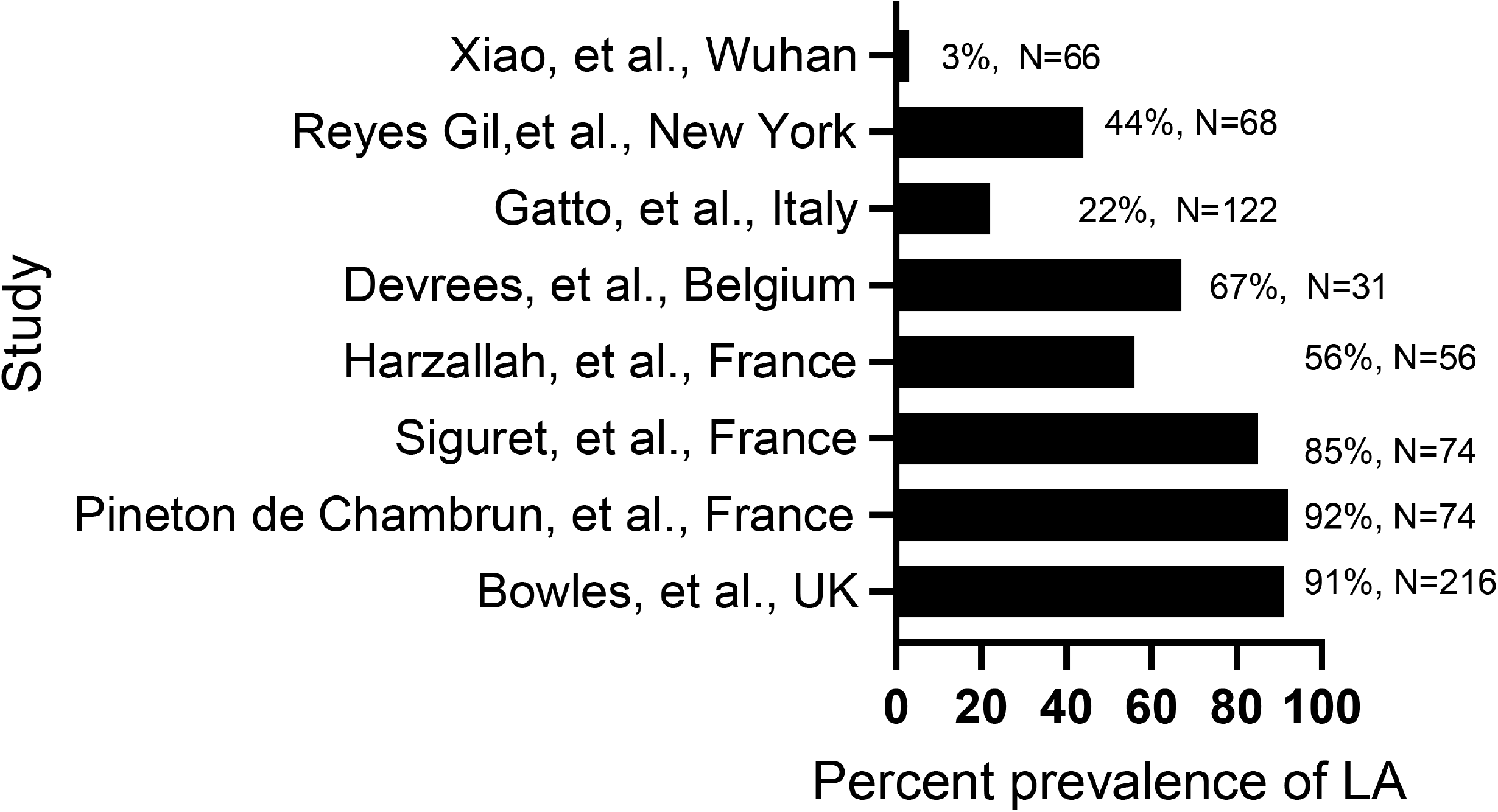
The percent prevalence of Lupus anticoagulant (LA) in COVID-19 patient populations: N= number of patients, Ref= Reference.

### Antiphospholipid positivity in patients with COVID-19

Studies from Europe and the United States have shown a very high prevalence of LA in patients with COVID-19, ranging from 19% to 92% (**Table 1, Figure 1**). The prevalence of LA was significantly higher in COVID-19 patients than in non-COVID-19 controls, for example, 44% vs. 22% in New York, and ∼19% vs. 2% in UK (9, 10). The prevalence of LA was also higher in Italian COVID-19 patients than in patients with other autoimmune rheumatic diseases (22% vs. 15%), but lower than in patients with primary APS (56%) (11). Contrary to these reports, patients from China were reported to have a much lower prevalence of LA – 3% of 66 and 5% of 19 patients in Wuhan and 0 of 13 in Beijing (12, 13). However, Chinese patients had a much higher prevalence of aβ2GP1 and aCL antibodies (47%-53%). These patients were tested at later time points (∼29-39 days) after disease onset than in most European and American studies. Consistently, aCL/aβ2GP1 positive French patients tended to have been tested longer after COVID-19 symptoms started than the negative patients (14). Furthermore, 10 Chinese patients who were aPL negative at an early time point after disease onset became aPL positive later (12), and a French patient with isolated IgM aCL seroconverted to IgG aCL and aβ_2_GP1 antibodies (14).

IgA aβ2GP1, IgA aCL, and IgG aβ2GP1 antibodies were the most prevalent aPL in Chinese patients, with many of them positive for multiple isotypes (12, 13). IgG aβ2GP1 antibodies were also most frequent in a report from Italy(15). Reports from Michigan and Mexico also showed a high prevalence of the non-criteria anti-phosphatidylserine/prothrombin antibodies (16, 17). IgA antibodies were not tested in many European and American studies.

As a whole, reports from around the world appear to suggest a high prevalence of LA at early time points after COVID-19 infection, whereas aPL antibodies emerge later in the disease course. This suggests that COVID-19 patients who have a longer disease duration are more likely to have aPL antibodies.

### Antiphospholipid positivity in relation to thrombosis in COVID-19 patients

An early report described three COVID-19 patients with multi-vessel brain infarction, acro-ischemia, and IgA/IgG aCL and aβ2GP1 antibodies (4). This was followed by a flurry of reports seeking a relation between aPL antibodies and thrombosis in COVID-19 (**Table 2** and **Appendix Table 2**).

**Table 2.** Reports of antiphospholipid assays in patients with COVID-19 and thrombosis

| Location [Ref.] | Patient cohort | Thrombotic manifestations | Relationship with aPL positivity |
| --- | --- | --- | --- |
| Wuhan, China (4) | 3 critically ill patients with COVID-19 | Multiple cerebral infarctions, acute limb ischemia, acro-ischemia | 3/3 + for multiple aPL. All had detectable IgA aCL and aβ2GP1 |
| Wuhan, China (12) | 66 critically ill patients with COVID-19 | Arterial in 6 (9%) patients (cerebral infarction 5, myocardial infarction 1), venous in 19 (29%) (large veins 2, distal veins 17) | Arterial and large vein thrombosis seen only in patients with multiple aPL at medium/high levels.<br>Cerebral infarction in aPL– vs. multiple aPL+ patients (0 vs. 33%, p 0.02). |
| Wuhan, China (13) | 19 ICU patients with COVID-19; all on anticoagulation | 12 thrombotic events in 9 patients (47%): 4 cerebral infarction, 7 acro-ischemia, and 1 internal jugular vein thrombosis. | 9/10 aPL+ vs. 0/9 aPL– patients had a thrombotic event.<br>All 4 patients with cerebral infarction had aPL of multiple isotypes incl. IgA aCL and aβ2GP1. |
| Italy (11) | 53 hospitalized patients and 69 in home quarantine; 89% on anticoagulation | 18/46 (39%) hospitalized patients had thrombosis, incl. 17 VTE and 1 stroke. No thrombosis in quarantined patients. | 50% of 53 hospitalized patients had at least 1 aPL (LA 17%, IgG/IgM/IgA aCL/aβ2GP1 ranged 4%-19%).<br>10/41 (24%) aPL+ vs. 8/65 (12%) aPL– patients had thrombosis (p 0.09). |
| Italy (15) | 122 severe or critical patients; 98% anticoagulated | 16 thrombotic events (13%), 8 each of venous and arterial. | No significant association between thrombosis and aPL (IgG/IgM aCL and IgG/IgM/IgA aβ2GP1 ranged 6%-16%). |
| France (14) | 74 mechanically ventilated patients on anticoagulation | 28 patients (38%) had a thrombosis, incl. 26 DVT, 4 PE, 1 stroke and 1 extensive venous catheter thrombosis. | IgG/IgM aCL and/or IgG aβ2GP1 + in 18% patients with thrombosis and 9% without thrombosis (p NS).<br>LA+ in 82% of 28 patients with thrombosis vs. 87% of 46 without it. |
| France (18) | 25 critically ill patients | PE in 6 patients | LA+ in 92% patients, aCL in 52%, Aβ2GP1 in 12%<br>All 6 patients who had PE were aPL+, of whom 4 were double + |
| Spain (19) | 24 of 785 COVID-19 patients who had VTE (3%) | DVT alone 9 (38%), PE alone 11 (46%), DVT+PE 4 (17%), arterial thrombosis 0 | 2 patients (8.3%) with weakly positive IgM aCL and aβ2GP1. None with IgG aPL |
| Belgium (20) | 31 ICU patients with COVID-19 | 12 events - CVC thrombosis 4, DVT 2, ECMO clotting 3, dialysis circuit 2, stroke 1 | 7/9 (78%) patients with thrombosis vs. 16/22 (73%) without thrombosis had at least 1 aPL. |
| UK (21) | 4 COVID-19 with stroke | Stroke (4/4), PE (1/4) | LA+ in 4/4 |
| Mexico (17) | 21 severe or critical patients; 18 anticoagulated | PE in 2/21 patients | 12/21 + for at least 1 aPL.<br>VTE in 2/12 aPL+ vs. 0/9 aPL–. |
| Michigan, USA (16) | 172 hospitalized patients; most on anticoagulation. | 11 patients with thrombosis: arterial 2 (1%), venous 8 (5%), both 1 (0.6%). | Any aPL + in 89 (52%) patients.<br>6/11 (55%) patients with thrombosis were aPL+. |
| New York, USA (10) | 68 patients who had LA test orders and PCR+ for SARS-COV-2 | 8 arterial events (incl. 2 stroke) and 11 VTE (incl. 3 PE) in LA+ patients vs. all VTE events in LA– patients | Thrombosis in 63% of 30 LA+ vs. 34% of 38 LA– patients (p <0.03).<br>LA independently associated with thrombosis: OR 4.4 (95% CI, 1.5-14.6), p 0.01. |
| UAE (42) | 22 patients with ischemic stroke | 22 acute ischemic stroke | Of 6 patients tested, 1 was +, Specific antibody not mentioned. |
| Brazil (43) | 5 patients with mild infection | All 5 with acute PE | 4/5 positive for LAC, 2/5 aCL IgM, 1/5 Aβ2 IgM |

+, positive; –, negative; aβ2GP1, anti-β2 glycoprotein 1 antibody; aCL, anti-cardiolipin antibody; aPL, antiphospholipid antibody; CVC, central venous catheter; CVD, cardiovascular disease; DM, diabetes mellitus; dRVVT, diluted Russell viper venom test; DVT, deep vein thrombosis; HTN, hypertension; incl., including; ICU, intensive care unit; LA, lupus anticoagulant; OR, odds ratio; PE, pulmonary embolism; VTE, venous thromboembolism

In 66 COVID-19 patients from Wuhan, 38% developed arterial or venous thrombosis, and nearly half of all patients were positive for aPL antibodies (12). When stratified based on positivity for aPL antibodies, patients with medium/high positivity for multiple aPL antibodies showed a significantly higher incidence of cerebral infarction compared to patients with negative aPL antibodies (0 vs. 33.3%, p 0.023). In another report from Wuhan, 9 of 10 patients who were aPL positive as compared to 0 of 9 aPL-negative patients developed thrombosis; all patients were already receiving anticoagulation(13). All four patients who developed cerebral infarction had multiple isotypes of aPL antibodies detectable.

Such a tight link between the presence of aPL antibodies and thrombosis in COVID-19 patients was not apparent in reports from Europe. Nonetheless, 24% of Italian patients with at least one aPL developed thrombosis versus 12% of patients who were aPL negative (p 0.09) (11). Thrombotic events in this report were almost exclusively venous, and 50% of all patients were positive for at least one aPL. In another report from Italy, 13% of patients had a thrombotic event including 8 each of arterial and venous thrombosis and 6%-16% had various aPL, but there was no significant association between thrombosis and aPL (15). Venous events, namely DVT and pulmonary embolism (PE), were also frequent in French patients, and aPL antibody positivity tended to be more frequent in patients with thrombosis than in those without it, but the differences were not statistically significant (14, 18). No association with aPL was found in patients from Spain and Belgium who had predominantly venous thrombosis (19, 20). Likewise, a Michigan study found no significant association between aPL positivity and thrombosis in patients who received a stricter thromboprophylaxis and had a low frequency of thrombotic complications (6%) (16). Overall, most reports from Europe did not support a significant relationship between aPL positivity and thrombosis in patients with COVID-19. Most of these studies had a relatively small number of thrombotic events and did not stratify them into arterial and venous events separately to ascertain specific relations with aPL antibodies.

In a New York cohort of 60 patients with COVID-19 who had 32 thrombotic events including 8 arterial and 24 venous, LA was found to be independently associated with thrombosis (10). All of the arterial thrombotic events and 11 venous thromboembolism occurred in LA-positive patients as compared to LA-negative patients who had only venous events. All 4 patients with COVID-19 and stroke in a UK report were also positive for LA (21). Overall, these reports suggest a potential link between aPL antibodies and thrombosis in COVID-19.

### Antiphospholipid antibodies and individual manifestations of APS in patients with COVID-19

Next, to seek any potential links between aPL and individual APS manifestations in COVID-19, we compared the frequency of aPL in patients with specific manifestations in COVID-19 to the general population, the latter as reported in a systematic review of 120 full-text papers (22). The overall aPL frequency in this analysis was estimated to be 13.5% for stroke, 11% for myocardial infarction, 9.5% for DVT, and 6% for pregnancy morbidity in the general population.

Of 438 COVID-19 patients with ischemic stroke, 65 were tested for aPL antibodies, and 38 (58.4%) were found to be positive (**Table 3**). Compared to the overall frequency of aPL in stroke patients in the general population, which was 13.5% (interquartile range 6.8-23.3) in all 31 studies and 7.5% (5.3-13.8) in 7 prospective studies(22), COVID-19 patients with stroke had a significantly higher frequency of aPL (p <0.0001, **Figure 2**). This profound difference in the prevalence of aPL antibodies in stroke patients with and without SARS-CoV-2 infection suggests a link between ischemic stroke and aPL antibodies in COVID-19, with a caveat that the two groups of patients are different in terms of comorbidities, age, state of health, and other demographic factors.

**Table 3.**
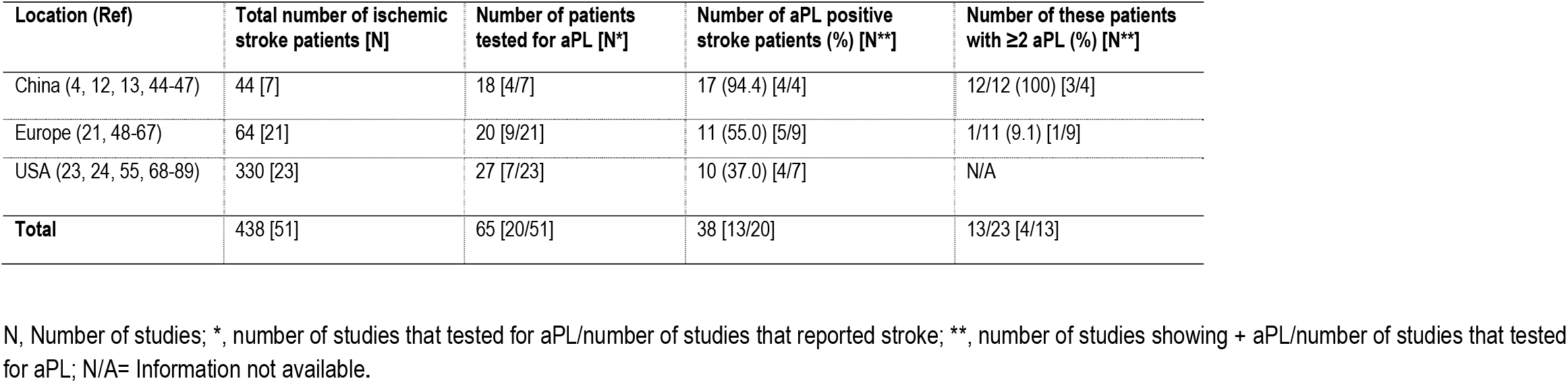
Cerebral infarction/stroke in patients with COVID-19

| Location (Ref) | Total number of ischemic stroke patients [N] | Number of patients tested for aPL [N*] | Number of aPL positive stroke patients (%) [N**] | Number of these patients with $\geq 2$ aPL (%) [N**] |
| --- | --- | --- | --- | --- |
| China (4, 12, 13, 44-47) | 44 [7] | 18 [4/7] | 17 (94.4) [4/4] | 12/12 (100) [3/4] |
| Europe (21, 48-67) | 64 [21] | 20 [9/21] | 11 (55.0) [5/9] | 1/11 (9.1) [1/9] |
| USA (23, 24, 55, 68-89) | 330 [23] | 27 [7/23] | 10 (37.0) [4/7] | N/A |
| **Total** | 438 [51] | 65 [20/51] | 38 [13/20] | 13/23 [4/13] |

N, Number of studies; \*, number of studies that tested for aPL/number of studies that reported stroke; \*\*, number of studies showing + aPL/number of studies that tested for aPL; N/A= Information not available.

**Figure 2.**
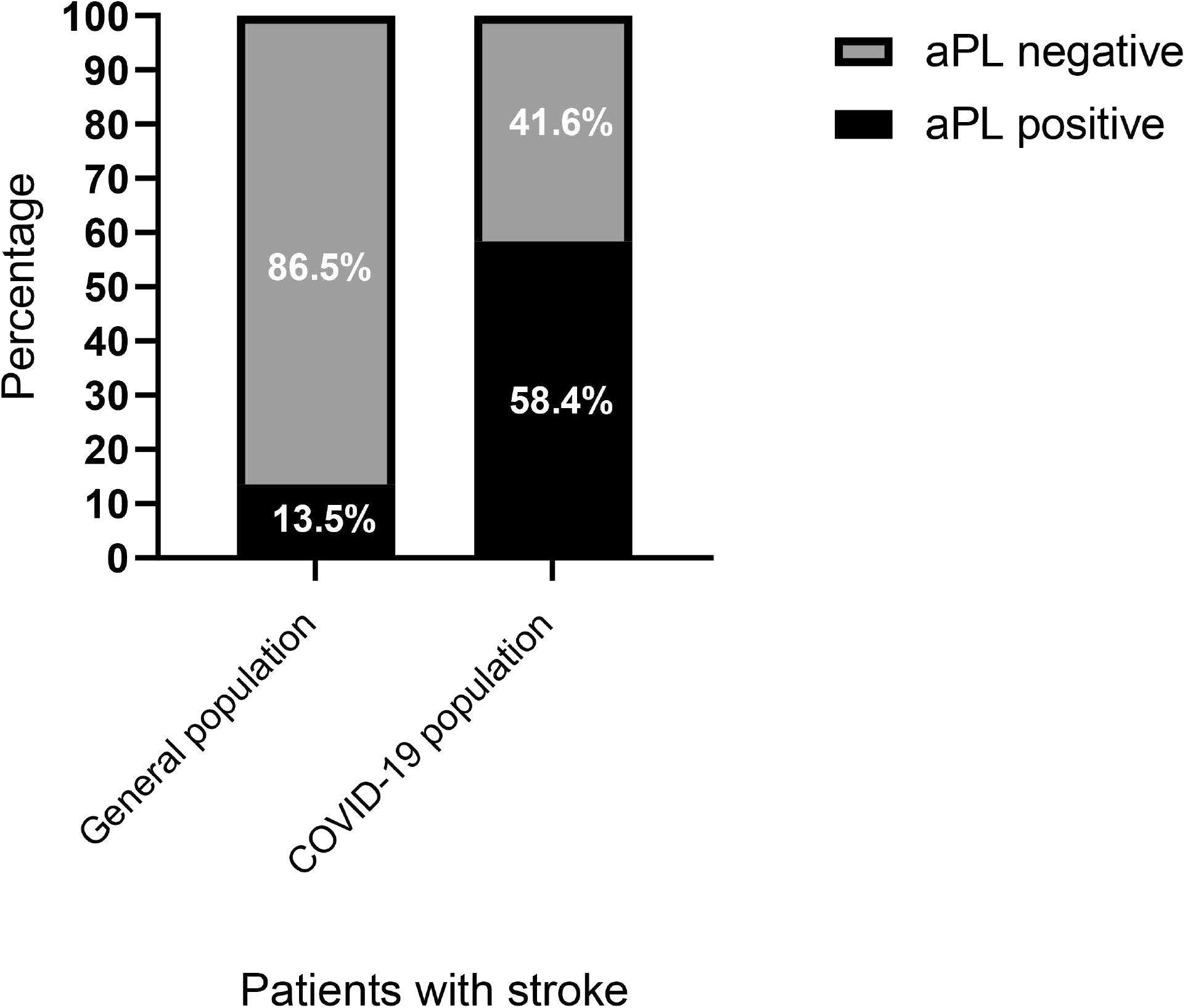
The percent prevalence of aPL in stroke patients with COVID-19 as compared to the general population. At least one aPL assay was positive in 38 of 65 (58.4%) patients with COVID-19 and stroke. The average age in these patient series was 68.0, 66.6, 65.2, 65.0, 64.0, 61.9, and 47.6 years. The overall frequency of aPL in stroke patients in general population was 13.5% (643/4765) in a recent review of 31 studies (22). The age of patients in these 31 studies ranged from 2 years to 88 years. The differences in age distribution, comorbidities, and overall health status are major limitation of our analysis. *p <0.0001, Fisher’s exact test.

There were notable differences in the prevalence of aPL antibodies in COVID-9 patients with stroke from different continents: 94.4% in Chinese patients with ischemic stroke, 55% in European patients, and 37% in American patients (**Table 3**).

In addition to stroke, arterial thrombosis in SARS-CoV-2 infection has manifested with myocardial infarction, acute limb ischemia, and thrombosis of large arteries (23, 24). Thus far, there are only a few reports of aPL antibodies in patients with myocardial infarction and acute limb ischemia (**Table 2, Appendix Table 2**).

Venous thrombosis in patients infected with SARS-CoV-2 has predominantly manifested as DVT and PE and occasionally as cerebral venous thrombosis and portal vein thrombosis. Among 150 COVID-19 patients referred to intensive care units of a French tertiary hospital, 64 thrombotic complications occurred, including PE (16.7%) (2). Notably, 87.7% of 57 patients suspected to have a coagulation disorder and tested had a positive LA. This frequency of aPL in venous thrombosis in COVID-19 patients appears much higher (27/87=31%) than aPL frequency in venous thrombosis patients in the general population (9.5%) (22). However, the testing and reporting of aPL are inadequate at this time for most of 648 patients with PE and 430 patients with DVT from around the world to make statistical comparisons.

Pregnancy loss is also a major manifestation of APS. Of 458 pregnancies in hospitalized patients with COVID-19 in the US, pregnancy loss occurred in 10 (2.2%) including four that occurred within 20 weeks of gestation (25). There have also been case reports of 1^st^ and 2^nd^-trimester miscarriages in asymptomatic or minimally symptomatic women (26). We have not found studies of aPL in relation to pregnancy outcome.

Thrombocytopenia occurs in 22%-42% of patients with APS. It has been reported in 5% to 54% of patients with COVID-19, with the incidence varying according to disease severity (27). It has also been reported in many asymptomatic patients. However, its association with aPL has not been explored in COVID-19.

Livedo reticularis-like vascular lesions, the most common skin manifestation of APS, have been reported in 2%-5% of patients with COVID-19 (28). Its association with aPL antibodies has not been explored.

Akin to reports in patients with APS, higher levels of aPL in COVID-19 patients were associated with neutrophil hyperactivity including the release of neutrophil extracellular traps (NET), and IgG fractions isolated from patients with COVID-19 promoted NET release from control neutrophils (16).

### Pathogenic significance of antiphospholipid antibodies

A report from Belgium found single LA positivity during the acute phase of COVID-19 infection, but not when tested one month later. This led them to suggest that COVID-19, like other viral infections, causes a transient production of aPL antibodies in critically ill patients, which may be of no clinical significance (6, 20). However, other studies detected aPL antibodies more than a month after the onset of symptoms, and many patients who were aPL negative at an early time point became positive later (12, 15). In Chinese studies, all symptomatic thrombotic events occurred in aPL positive patients (4, 12, 13). Higher levels of aPL were also associated with more severe respiratory disease and lower glomerular filtration rate in COVID-19 patients(16). Importantly, similar to studies in APS (29), injection of IgG fractions from COVID-19 patients into mice accelerated venous thrombosis (16). Collectively, these studies suggest that aPL are potentially thrombogenic in patients with COVID-19.

## Discussion

Viral infections are known to elicit the production of aPL antibodies, but the antibodies are generally transient and rarely cause thrombosis except for HIV and HCV that mainly induce IgG aCL antibodies (6). Thromboembolic events in patients with HCV were predominantly in patients who had aCL antibodies and were negative for LA and aβ2GP1(7). This is different from the aPL profile reported in COVID-19. The presence of LA was independently associated with thrombosis in patients with COVID-19 in some, but not in all studies. In COVID-19 patients in Wuhan, positivity for multiple aPL, including aCL and aβ2GP1 antibodies, of multiple isotypes at medium-to-high titers were significantly associated with arterial thrombosis (12, 13), which is consistent with the literature on APS that double and triple aPL positivity is associated with the increased risk for thrombosis (30).

IgA aCL, IgA aβ2GP1, and IgG aβ2GP1 antibodies were the most common aPL in COVID-19 patients in Wuhan (12). There was a striking association between the presence of these antibodies and cerebral infarction. The latter has important implications, as IgA aβ2GPI has been suggested as a risk factor for acute myocardial infarction and acute cerebral ischemia (31). Other studies have also linked IgA aPL antibodies to a higher risk of thrombosis (32). It is postulated that the infection of the respiratory and gastrointestinal mucosa by SARS-CoV-2 may elicit the IgA antibody production, which could go on to cause thrombosis (12). These observations warrant that IgA aPL, although not part of the diagnostic criteria for APS, be uniformly tested in addition to IgG/M aPL in patients with acute arterial ischemic events and COVID-19.

The significantly higher aPL antibody positive rate in China compared to the USA and Europe could be due to various reasons. Chinese studies tested IgA antibodies, while patients in Europe and the US were mostly not tested for them. Chinese patients with stroke could also have had SARS-CoV-2 infection for a longer duration. It is also possible that APS is an important mechanism of thrombosis in severely ill SARS-CoV-2 patients, which had not been (at the time of this study) adequately tested in the US and Europe.

Our analysis has several limitations. First, the Sapporo classification criteria for APS requires the aPL to be measured twice over a period of 12 weeks to establish that the levels are not transient (33). In this regard, most studies have tested aPL in hospitalized patients with SARS-CoV-2 infection within days of the onset of symptoms. Therefore, follow up studies are required to truly understand whether the patient developed APS or simply had a transient elevation of aPL.

Second, the guidelines from the International Society on Thrombosis and Hemostasis state that anticoagulation must be discontinued for 12 hours before the testing for LA or the testing must be done before anticoagulation is started (34). Since critically ill patients are often anticoagulated, it is important to interpret a positive LA result with caution in this setting. Elevated levels of the acute phase reactant C-reactive protein, as commonly seen in patients hospitalized with COVID-19, have also been shown to interfere with reagents commonly used to test LA (35). Antibody assays for aCL and aβ2GP1 antibodies also suffer from a lack of standardization and the presence of inter-laboratory variation, which can influence the interpretation of tests with borderline results (36).

Third, many patients with COVID-19 in most reports analyzed had comorbidities including diabetes and hypertension, which could contribute to a pro-thrombotic milieu. Other complications, including heparin-induced thrombocytopenia, disseminated intravascular coagulation, and thrombotic microangiopathy, could also be causes of thrombosis in these critically ill patients. We also acknowledge the risk of bias including, but not limited to, possible poor/inadequate reporting in individual articles, potential lack of external validity, potential selective outcome reporting in individual articles, evidence selection bias due to non-publication, new information published after our cut-off date, and articles in non-English languages,

Finally, it has been suggested that catastrophic APS could be a possible explanation of the cytokine storm in some patients with severe COVID-19. Catastrophic APS is a fulminant form of APS that presents with widespread small vessel thrombosis. However, unlike catastrophic APS that manifests predominantly with intra-abdominal thrombosis, patients who died from COVID-19 had a predominance of pulmonary microvascular thrombosis (37).

Studies in APS suggest a direct pathogenic role of aPL antibodies, as the experimental transfer of polyclonal IgG antibodies against domain I of β2 glycoprotein from a patient with APS into mice induced thrombosis in blood vessels (29). Consistently, injection of IgG fractions from COVID-19 patients who have aPL antibodies into mice accelerated venous thrombosis (16). The current working model of APS pathogenesis invokes a “two-hit hypothesis”, whereby the presence of aPL antibodies (the first-hit) would predispose to thrombosis upon exposure to a second hit such as infection or local tissue damage (38). Antiphospholipid antibodies are postulated to cause thrombosis via activation of endothelial cells, neutrophils, monocytes, platelets, and the complement cascade (39), which are similarly activated in COVID-19 infection likely due to the cytokine storm that occurs in some patients with COVID-19 (40). Taken together, it would be reasonable to postulate that the presence of aPL antibodies along with activation of endothelial and other immune cells could underlie the serious vascular complications in patients with COVID-19.

There are wide-ranging implications if it is proven that aPL antibodies are thrombogenic in COVID-19. For one, testing of aPL could be recommended for critically ill patients if it is proven that aPL positivity is a harbinger of future thrombotic events. In addition, the patients who have detectable aPL antibodies for 12 weeks or more can be diagnosed with APS and thus become candidates for long-term anticoagulation, thereby decreasing their long-term risk of vascular events.

## CONCLUSIONS

In conclusion, a substantial proportion of patients with COVID-19 exhibit aPL positivity. Follow-up testing is needed to identify whether these antibodies persist and pose a thrombosis risk in the future. Experimental transfer studies showing venous thrombosis in mice injected with human IgG from COVID-19 patients with aPL suggest a potential pathogenicity of these antibodies (16), however, more studies are needed to establish a causal relation between aPL positivity and thrombosis in COVID-19.

## Data Availability

Data are available upon reasonable request.

