## Supplemental Material for "Vascular Thrombosis in COVID-19: A Potential Association with Antiphospholipid Antibodies"

### CONTENTS

1. Appendix Figure 1. Search results in our systematic review.
2. Appendix Table 1. Quality of evidence.
3. Appendix Table 2. Case Reports of patients with COVID-19 and thrombosis who had antiphospholipid assays. This Table includes reports of single and two cases with thrombosis and antiphospholipid testing.
4. Appendix Table 3. Comorbidities in patients with COVID-19 and thrombosis who had antiphospholipid testing.
5. Supplementary References

**Appendix Figure 1**. **Search results in our systematic review.**

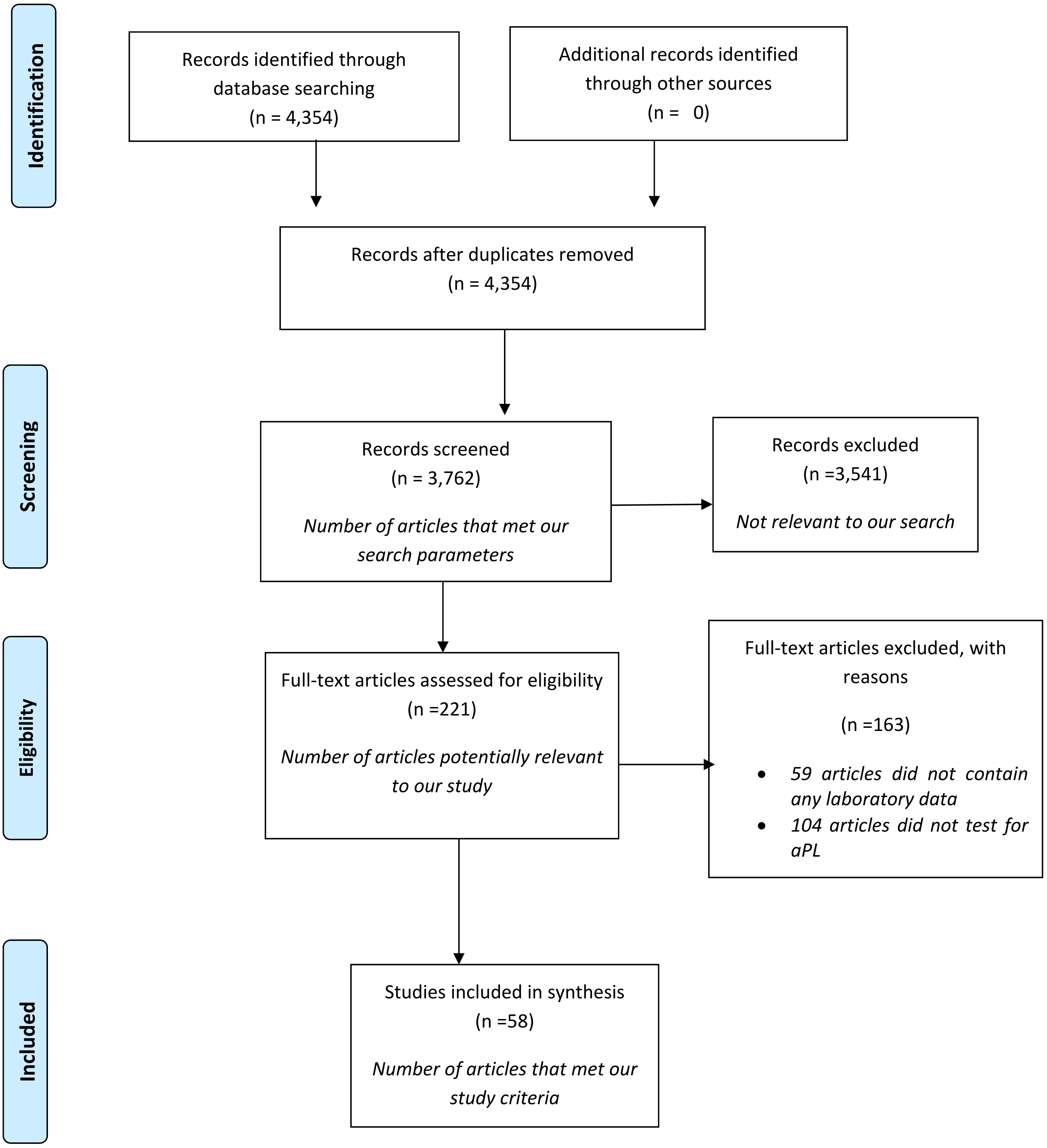

### Appendix Table 1. Quality of Evidence

| Study | Type of evidence | Quality of evidence |
|---|---|---|
| *Al-Ani, et al.* | Systematic review with meta-analysis | 1 |
| *Helms, et al.* | Prospective cohort study | 3 |
| *Wichmann, et al.* | Prospective cohort study | 3 |
| *Zhang, et al.* | Case series without intervention | 4 |
| *Bowles, et al.* | Retrospective cohort study | 3 |
| *Reyes Gil, et al.* | Retrospective cohort study | 3 |
| *Gatto, et al.* | Cross-sectional study | 4 |
| *Xiao, et.al* | Cross-sectional study | 4 |
| *Zhang et al.* | Cross-sectional study | 4 |
| *Siguret, et al.* | Cross sectional study | 4 |
| *Borghi, et al.* | Cross-sectional study (Not peer-reviewed) | 4 |
| *Zuo, et al.* | Cross-sectional study (Not peer-reviewed) | 4 |
| *Pineton De Chambrun, et al.* | Cross sectional study | 4 |
| *Galeano-Valle, et al.* | Cross sectional study | 4 |
| *Devreese, et al.* | Cross-sectional study | 4 |
| *Beyrouti, et al.* | Case series without intervention | 4 |
| *Etkin, et al.* | Cross sectional study | 4 |
| *Kashi, et al.* | Case series without intervention | 4 |
| *Delahoy, et al.* | Cross sectional study | 4 |
| *Wong, et al.* | Case series without intervention | 4 |
| *Freeman, et al.* | Case series without intervention | 4 |
| *Elsoukkary, et al.* | Case series without intervention | 4 |
| *Harzallah, et al.* | Cross sectional study | 4 |
| *Khan, et al.* | Case series without intervention | 4 |
| *Vechi, et al.* | Case series without intervention | 4 |
| *Mao, et al.* | Case series without intervention | 4 |
| *Fan, et al.* | Case series without intervention | 4 |
| *Xiong, et al.* | Retrospective cohort study | 3 |
| Barrios-López*, et al.* | Case series without intervention | 4 |
| *Basi, et al.* | Case report | 5 |
| *Bigliardi, et al.* | Case report | 5 |
| *Bonardel, et al.* | Case report | 5 |
| *D'Anna, et al.* | Case series without intervention | 4 |
| *Diaz-Perez, et al.* | Case series without intervention | 4 |
| *Duroi, et al.* | Case report | 5 |
| *Fara, et al.* | Case report | 5 |
| *Gemcioglu, et al.* | Case report | 5 |
| González-Pinto*, et al.* | Case report | 5 |
| *Helms, et al.* | Case series without intervention | 4 |
| *Immovilli, et al.* | Case series without intervention | 4 |
| *Klok, et al.* | Cross sectional study | 4 |
| *Lapergue, et al.* | Case series without intervention | 4 |
| *Lodigiani, et al.* | Retrospective cohort study | 3 |
| *Mohamed, et al.* | Case report | 5 |
| *Morassi, et al.* | Case series without intervention | 4 |
| *Roy, et al.* | Case series without intervention | 4 |
| TunÇ*, et al.* | Case series without intervention | 4 |
| *Zayet, et al.* | Case series without intervention | 4 |
| *Avula, et al.* | Case series without intervention | 4 |
| *Deliwala, et al.* | Case report | 5 |
| *Elshereye, et al.* | Case report | 5 |
| *Esenwa, et al.* | Case series without intervention | 4 |
| *Garg, et al.* | Case report | 5 |
| *Goldberg, et al.* | Case report | 5 |

| | | |
|---|---|---|
| *Gunasekaran, et al.* | Case report | 5 |
| *Jillella, et al.* | Case series without intervention | 4 |
| *Kariyanna, et al.* | Case report | 5 |
| *Katz, et al.* | Case series without intervention | 4 |
| *Kihira, et al.* | Case-control study | 3 |
| *Kvernland, et al.* | Retrospective cohort study | 3 |
| *Xia, et al.* | Retrospective cohort study | 3 |
| *Mahboob, et al.* | Case report | 5 |
| *Majidi, et al.* | Case series without intervention | 4 |
| *Oxley, et al.* | Case series without intervention | 4 |
| *Reddy, et al.* | Case series without intervention | 4 |
| *Rothstein, et al.* | Case series without intervention | 4 |
| *Valderrama, et al.* | Case report | 5 |
| *Yaghi, et al.* | Retrospective cohort study | 3 |
| *Zahid, et al.* | Case report | 5 |

**Supplementary Table**

| Author and Citation Number | Type of Evidence | Quality of Evidence |
|---|---|---|
| *Hossri, et.al.* | Case series without intervention | 4 |
| *Dumitrascu, et.al.* | Case report | 5 |
| *Shams, et.al.* | Case report | 5 |
| *Zendjebil, et.al.* | Case report | 5 |
| *Soltani, et.al.* | Case report | 5 |
| *Gomez-Arbelaez, et.al.* | Case series without intervention | 4 |
| *Woehl, et.al.* | Case series without intervention | 4 |
| *Nassabein, et.al.* | Case report | 5 |
| *Mosbahi, et.al.* | Case report | 5 |
| *Veyre, et.al.* | Case report | 5 |
| *Singh, et.al.* | Case series without intervention | 4 |
| *Warrior, et.al.* | Case report | 5 |
| *Renaud-Picard, et.al.* | Case report | 5 |
| *Garaci, et.al.* | Case report | 5 |
| *Klein, et.al.* | Case report | 5 |
| *Roy-Gash, et.al.* | Case report | 5 |
| *Bolaji, et.al.* | Case report | 5 |
| *Hemasian, et.al.* | Case report | 5 |
| *Alharthy, et.al.* | Case series without intervention | 4 |
| *Marsico, et.al.* | Case series without intervention | 4 |
| *Akiyama, et.al.* | Case report | 5 |
| *Ofosu, et.al.* | Case report | 5 |
| *Franco-Moreno, et.al.* | Case report | 5 |
| *Del Hoyo, et.al.* | Case report | 5 |
| *Pang, et.al.* | Case report | 5 |
| *Fan, et.al.* | Case report | 5 |
| *Veyseh, et.al.* | Case report | 5 |
| *Sung, et.al.* | Case report | 5 |
| *Davoodi, et.al.* | Case report | 5 |
| *Nauka, et.al.* | Case report | 5 |

**Appendix Table 2**. **Case Reports of patients with COVID-19 and thrombosis who had antiphospholipid assays.**

This Table includes reports of single and two cases with thrombosis and aPL testing.

| Location (Ref) | Patient cohort | Thrombotic manifestations | aPL test |
|---|---|---|---|
| **Arterial thrombotic events** | | | |
| USA (1) | 2 critically ill patients | Multiple cerebral infarction and splenic infarct (patient 1), Acute limb ischemia (patient 2) | IgM and IgG aCL + in both |
| USA (2) | 1 patient (48/M) | Ophthalmic artery occlusion | – |
| Qatar (3) | 1 patient (28/M) | Coronary thrombosis | aCL – |
| France (4) | 1 patient (42/M) | Myocardial infarction | – |
| Canada (5) | 1 patient (63/F) | Myocardial infarction | – |
| Spain (6) | 4 patients (50-76 years) | Aortic thrombosis | 2/4 patients were tested; both – |
| France (7) | 4 patients (64-78 years) | Aortic thrombosis | 2/4 patients tested; both – |
| Canada (8) | 1 patient (66/F) | Aortic thrombosis | – |
| Switzerland (9) | 1 patient (58/M) | Aortic thrombosis | Thrombophilia workup –, exact aPL status unclear |
| France (10) | 1 patient (24/M) | Femoral artery thrombosis | Negative |
| USA (11) | 3 patients (71/F, 70/M, 70/F) | Acute limb ischemia | Only 2 were tested, both trace + for IgM aCL |
| USA (12) | 1 patient (59/F) | Acute limb ischemia | **IgM aCL +** |
| France (13) | 1 patient (31/M) | Acute limb ischemia | **LA +** |
| **Venous thrombotic events** | | | |
| Italy (14) | 1 patient (44/F) | Cerebral venous thrombosis | – |
| USA (15) | 1 patient (29/F) | Cerebral venous thrombosis | **aCL +** |
| France (16) | 1 patient (63/F) | Cerebral venous thrombosis | – |
| UK(17) | 1 patient (63/M) | Cerebral venous thrombosis | **LA low titer +** |
| Iran (18) | 1 patient (65/M) | Cerebral venous thrombosis | Thrombophilia workup –, aPL status unclear. |
| Saudi Arabia (19) | 2 patients- 50/F, 56/F | Acute pulmonary embolism | Thrombophilia workup –, aPL status unclear |
| Spain (20) | 2 patients- 32/M, 59/F | Pulmonary embolism | **LA +,** not specified for patient 2 |
| Japan(21) | 1 patient (56/M) | Pulmonary embolism | – |
| USA (22) | 1 patient (55/M) | Portal vein thrombosis | LA – |
| Spain (23) | 1 patient (27/M) | Portal vein thrombosis | – |
| Spain (24) | 1 patient (61/F) | Splanchnic vein thrombosis | **Low titer LA +** |
| Singapore (25) | 1 patient (30/M) | Superior mesenteric vein thrombosis | **LA +** |
| Singapore (26) | 1 patient (30/M) | Superior mesenteric vein thrombosis | **LA +** |
| USA(27) | 1 patient (52/F) | Gonadal vein thrombosis | LA – |
| USA (28) | 1 patient | Four extremity DVT | LA and IgM/IgG aCL + |
| Iran(29) | 1 patient (57/F) | DVT | aCL – |
| USA (30) | 1 patient (48/M) | DVT | – |

–, negative; +, positive; aCL, anti-cardiolipin antibody; aPL, antiphospholipid; CAD, coronary artery disease; DM, diabetes mellitus; DVT, deep vein thrombosis; F, female; HTN, hypertension; LA, lupus anticoagulant; M, male.

Overall, we found reports of positive aPL antibodies in all 8 patients with acute limb ischemia, 1 of 4 patients with myocardial infarction, none of 6 with aortic thrombosis, 2 of 5 patients with cerebral venous thrombosis, and all 3 patients with splanchnic/superior mesenteric venous thrombosis.

**Appendix Table 3. Comorbidities in patients with COVID-19 and thrombosis who had antiphospholipid testing.**

| Location [Ref.] | Patient cohort | Comorbidites |
|---|---|---|
| Wuhan, China (31) | 3 critically ill patients with COVID-19 | HTN (3/3), DM (2/3), stroke (2/3) |
| Wuhan, China (32) | 66 critically ill patients with COVID-19 | HTN (≈ 50%), DM (≈ 20%) |
| Wuhan, China (33) | 19 ICU patients with COVID-19; all on anticoagulation | 63% with comorbidities |
| Italy (34) | 53 hospitalized patients and 69 in home quarantine; 89% on anticoagulation | Not mentioned |
| Italy (35) | 122 severe or critical patients; 98% anticoagulated | 6 patients with thrombotic event in the past |
| France (36) | 74 mechanically ventilated patients on anticoagulation | DM and HTN in 43% of those with thrombosis and 39%-46% of those without it. |
| France (37) | 25 critically ill patients | Not mentioned |
| Spain (38) | 24 of 785 COVID-19 patients who had VTE (3%) | Not mentioned |
| Belgium (39) | 31 ICU patients with COVID-19 | 45% CVD, 25.8% DM, 12.9% obesity |
| UK (40) | 4 COVID-19 with stroke | HTN (3/4), DM (2/4) |
| Mexico (41) | 21 severe or critical patients; 18 anticoagulated | HTN 12 (57%), DM 8 (39%), obesity 7 (33%) |
| Michigan, USA (42) | 172 hospitalized patients; most on anticoagulation. | HTN (68%), DM (41%), obesity (51%), smoking (26%) |
| New York, USA (43) | 68 patients who had LA test orders and PCR+ for SARS-n-COV-2 at a tertiary hospital | Ventilation, mortality, and anticoagulation similar in LA+ vs. LA– patients |
| UAE (44) | 22 patients with ischemic stroke and COVID-19 | 36% DM, 32% HTN, 9% CVD, 9% dyslipidemia |
| Brazil (45) | 5 patients with mild infection | 1 DM, 1 obesity and HTN |
| USA (1) | Two critically ill patients with COVID-19 | Sickle cell trait (patient 1)<br>Dyslipidemia (patient 2) |
| USA (2) | 1 patient (48/M) | Obesity |
| Qatar (3) | 1 patient (28/M) | None |
| France (4) | 1 patient (42/M) | None |
| Canada (5) | 1 patient (63/F) | COPD |
| Spain (6) | 4 patients (50-76 years) | 1 each with HTN, and HTN+ dyslipidemia |
| France (7) | 4 patients (64-78 years) | 2 with HTN+ dyslipidemia, 1 with HTN |
| Canada (8) | 1 patient (66/F) | HTN, malignancy |
| Switzerland (9) | 1 patient (58/M) | Not mentioned |
| France (10) | 1 patient (24/M) | None |
| USA (11) | 3 patients (71/F, 70/M, 70/F) | 1 with dementia, Parkinson’s,<br>1 HTN,<br>1 DM + HTN |
| USA (12) | 1 patient (59/F) | HTN, hyperlipidemia |
| France (13) | 1 patient (31/M) | Cystic fibrosis, bilateral lung transplantation |
| Italy (14) | 1 patient (44/F) | None |
| USA (15) | 1 patient (29/F) | None |
| France (16) | 1 patient (63/F) | Not mentioned |

| | | |
|---|---|---|
| UK(17) | 1 patient (63/M) | DM |
| Iran (18) | 1 patient (65/M) | None |
| Saudi Arabia (19) | 2 patients- 50/F, 56/F | None |
| Spain (20) | 2 patients- 32/M, 59/F | None, HTN |
| Japan(21) | 1 patient (56/M) | None |
| USA (22) | 1 patient (55/M) | Hyperlipidemia |
| Spain (23) | 1 patient (27/M) | None |
| Spain (24) | 1 patient (61/F) | DM |
| Singapore (25) | 1 patient (30/M) | None |
| Singapore (26) | 1 patient (30/M) | None |
| USA(27) | 1 patient (52/F) | None |
| USA (28) | One patient | None |
| Iran(29) | 1 patient (57/F) | None |
| USA (30) | 1 patient (48/M) | CAD, renal donation |

+, positive; –, negative; aβ2GP1, anti-ß2 glycoprotein 1 antibody; aCL, anti-cardiolipin antibody; aPL, antiphospholipid antibody; CVC, central venous catheter; CVD, cardiovascular disease; DM, diabetes mellitus; dRVVT, diluted Russell viper venom test; DVT, deep vein thrombosis; HTN, hypertension; incl., including; ICU, intensive care unit; LA, lupus anticoagulant; OR, odds ratio; PE, pulmonary embolism; VTE, venous thromboembolism
